# Saliva cell type DNA methylation reference panel for epidemiology studies in children

**DOI:** 10.1101/2020.09.14.20191361

**Authors:** Lauren Y. M. Middleton, John Dou, Jonah Fisher, Jonathan A. Heiss, Vy K. Nguyen, Allan C. Just, Jessica Faul, Erin B. Ware, Colter Mitchell, Justin A. Colacino, Kelly M. Bakulski

## Abstract

Saliva is a widely used biological sample, especially in pediatric research, containing a heterogenous mixture of immune and epithelial cells. Associations of exposure or disease with saliva DNA methylation can be influenced by cell-type proportions. Here, we developed a saliva cell-type DNA methylation reference panel to estimate interindividual cell-type heterogeneity in whole saliva studies. Saliva was collected from 22 children (7-16 years) and sorted into immune and epithelial cells, using size exclusion filtration and magnetic bead sorting. DNA methylation was measured using the Illumina MethylationEPIC BeadChip. We assessed cell-type differences in DNA methylation profiles and tested for enriched biological pathways. Immune and epithelial cells differed at 164,793 (20.7%) DNA methylation sites (t-test p < 10^-8^). Immune cell hypomethylated sites mapped to genes enriched for immune pathways (p < 3.2 × 10^-5^). Epithelial cell hypomethylated sites were enriched for cornification (p = 5.2 × 10^-4^), a key process for hard palette formation. Saliva immune and epithelial cells have distinct DNA methylation profiles which can drive whole saliva DNA methylation measures. A primary saliva DNA methylation reference panel, easily implemented with an R package, will allow estimates of cell proportions from whole saliva samples and improve epigenetic epidemiology studies by accounting for measurement heterogeneity by cell-type proportions.

## Introduction

DNA methylation is an important mechanism regulating gene expression in both normal development and disease progression.^1^ During cellular differentiation, developmental genes are silenced and other cell-type-specific genes are activated via altered DNA methylation^2^ which results in a unique DNA methylation profile for each cell type.^3^ Disease processes or environmental exposures can also differentially alter DNA methylation patterns in cells and tissues.^4–6^ Epigenetic associations with exposures or disease outcomes are typically assessed in bulk tissues such as blood and saliva. Changes in tissue level DNA methylation profiles, however, could be caused by varying cellular responses. For example, a disease or exposure could shift average DNA methylation across all cells, shift the proportions of cell types, or shift DNA methylation in a particularly susceptible cell type.^7^ Bulk tissues are comprised of complex cell-type mixtures and have DNA methylation profiles that are vulnerable to alterations from diseases and exposures.

Accounting for cell-type proportions is essential in bulk tissue DNA methylation studies. Cell-type proportions can mediate or confound differences in DNA methylation associated with exposures or diseases. For example, initial widespread age-related DNA methylation associations in blood were later largely attributed to age-related differences in immune cell-type proportions.^8^ To estimate cell-type composition in bulk tissue using DNA methylation measures, cell-type-specific DNA methylation reference profiles are used. DNA methylation reference panels are available for immune cells in primary adult blood and cord blood, as well as epithelial and fibroblast cell types; however, a primary DNA methylation reference panel is not available for children.^9–11^ Saliva is a commonly used tissue as a non-invasive source of biological material in epigenetic epidemiology studies, particularly in children. A primary DNA methylation reference panel method for estimating cell-type proportions in saliva is needed.

Saliva is a commonly used tissue as a non-invasive source of biological material in epigenetic epidemiology studies, particularly in children. The use of saliva as a surrogate tissue in epigenetic epidemiology studies is increasing. A search of PubMed using the terms “saliva AND (epigenetics or DNA methylation) NOT chemistry” showed that between 1994 and 2019 there have been 213 papers published that used saliva as the source material for DNA methylation studies. Over this time period, the number of papers has trended upward. Saliva offers an easier collection strategy, particularly for vulnerable populations, and is noted as source of DNA methylation with similar quality to blood or other difficult to access tissues.^12–14^ One study found that they were able to obtain more DNA from saliva compared to blood and the data quality was high from both tissue types.^14^ The use of saliva for epigenetic studies is expected to continue to increase, which highlights the need for a saliva-specific cell-type investigation.

Saliva includes a heterogenous mixture of immune and epithelial human cells. Resident immune cells are present in oral tissues, and immune cells can leave the bloodstream and enter the oral cavity.^15,16^The keratinized epithelium is found in areas of the oral cavity that experience mechanical forces.^17^ Keratinocytes, a large (30-100µm diameter) epithelial cell covering the hard palette, undergo cornification as a mechanism of programmed cell death.^18^ Both immune and epithelial cells contribute to the overall DNA methylation profile of saliva and must be considered for the cell-type proportion estimation.^19^ Inter-individual differences in saliva cell composition may lead to wide variation in DNA methylation profiles, independent of exposure or disease status.^20^ Cell-type deconvolution methods estimate proportions of cell types from bulk tissue using differentially methylated sites in the genome.^21^ A saliva-based DNA methylation reference panel adapted for deconvolution would improve the biologic interpretability of existing and future epigenetic studies in saliva.

To assist epigenetic epidemiology studies, the objective of this study was to provide a DNA methylation reference panel for cell-type proportion estimation in children’s saliva. Our goals were: 1) Develop methods to enrich for cell types from children’s saliva; 2) Characterize and quantify differences in DNA methylation profiles of children’s saliva cell types; and 3) Apply our new cell-type reference panel to estimate cell proportions in whole saliva and compare our new cell-type reference panel to an existing method for estimating cell proportions.

## Materials and Methods

### Study sample and saliva collection

Children between the ages of 7 and 17 years were eligible for the current study. We recruited a convenience sample of 22 children from schools in Ann Arbor, Michigan. Parents were contacted via email. We obtained written, informed consent from a parent or guardian and verbal assent from the child. This study was approved by the University of Michigan Institutional Review Board (HUM00154853). We collected the demographic data (child’s sex, age, race, and whether the child was sick in the last 3 days) via written survey.

Prior to saliva collection, participants did not eat or drink for 30 minutes. Unstimulated saliva was collected into an empty 15mL tube (Falcon, CAT# 14-959-53A). Between 1.75 to 6.5mL of saliva were obtained per participant. Samples were stored at room temperature before processing, and storage time ranged from 1-18 hours. Six of these participants additionally provided saliva samples directly into Oragene kits (DNA Genotek, CAT# OG-250).

### Saliva processing & cell enrichment

The Oragene kit samples were mixed using a 1mL pipet and a 500µL aliquot was removed and stored on ice in a microcentrifuge tube (Corning, CAT# 3621) until DNA extraction. Saliva samples collected in Falcon tubes were processed into three components: composite “Whole” saliva, enriched “Epithelial” cells, and enriched “Immune” cells. The overall workflow depicting the isolation of the cell fractions is shown in **Figure 1a**. Saliva cells were washed by diluting the samples up to 14mL with DPBS (Gibco, CAT# 14190144) and centrifuging at 500g for 5 minutes at 4°C. The supernatant was removed, and the pellet was resuspended in 10mL DPBS. The samples were centrifuged at 350g for 5 minutes at 4°C, and the supernatant was removed.

**Figure 1.**
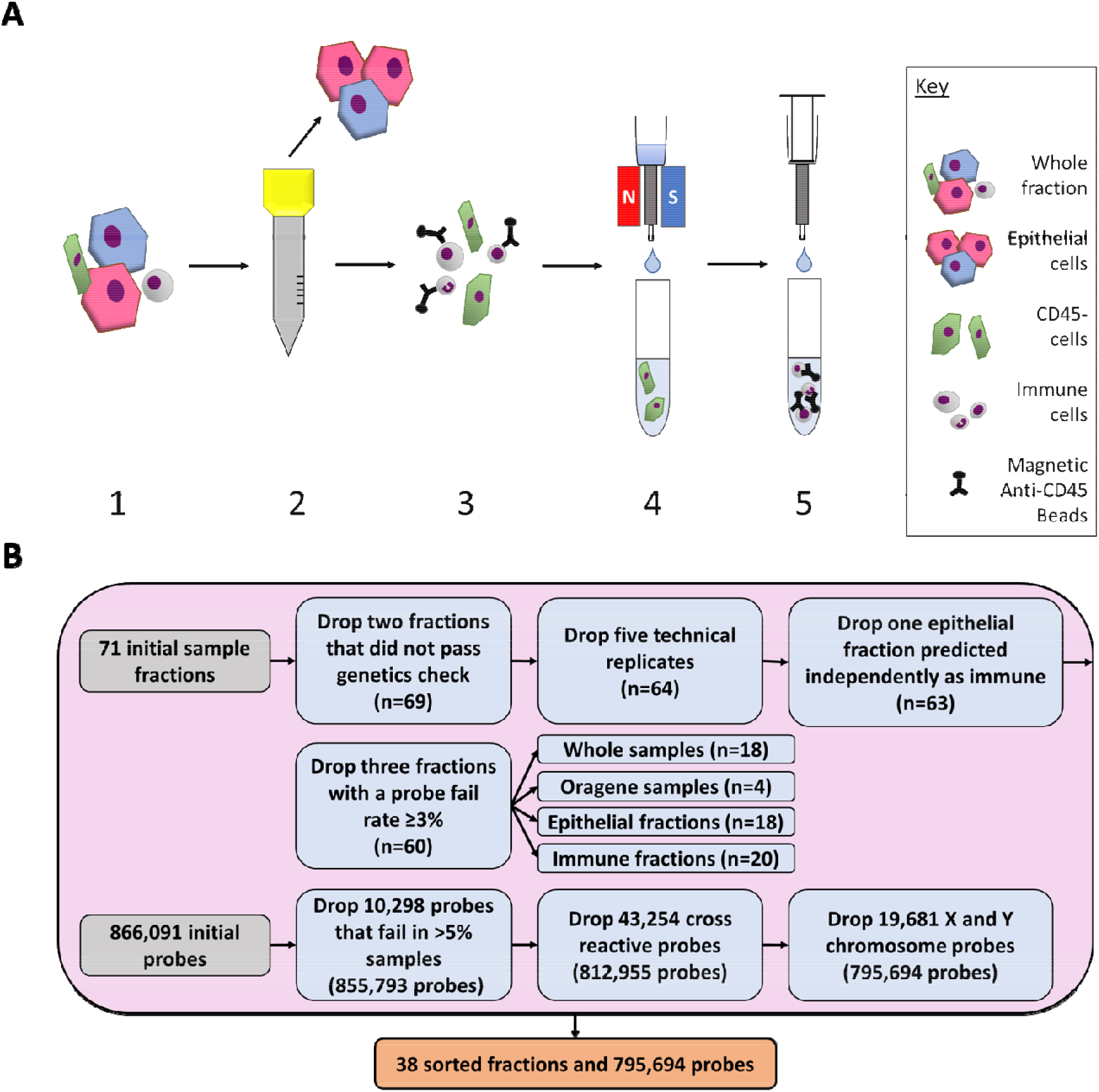
Diagram of experimental workflow. **A)** Saliva samples were sorted using size exclusion filtration and antibody-based magnetic bead methods. 1) Whole saliva samples were collected from participants. Samples were diluted and centrifuged. 2) The sample was passed through a 30µm filter. Cells captured on the filter were then rinsed into a separate collection tube. 3) The small cells that passed through the filter were mixed with CD45+ magnetic antibody dy beads to label the immune cells. 4) The sample was passed through a magnetized column which captured the immune cells in the column. Unlabeled cells flowed through into the collection tube. 5) The magnet was removed, and the immune cells and magnetic beads were eluted into a new collection tube. **B)** Quality control checks were used on the DNA methylation data from the whole samples, Oragene samples, epithelial fractions, and immune fractions. Samples were dropped before the probes. A total of 38 fractions and 795,694 probes were used in this analysis.

Fresh Bovine Serum Albumin (BSA) Rinsing Buffer was prepared each processing day with final concentrations of 0.5% BSA Fraction V (Gibco, CAT# 15260037), 2mM EDTA (Lonza, CAT# 51201), and DPBS. The BSA Rinsing Buffer sat on ice for a minimum of 10 minutes to degas. Cell pellets were resuspended in 3mL BSA Rinsing Buffer. The count and percent viability of cells were recorded using the LUNA-FL™ Dual Fluorescence Cell Counter (Logos Biosystems; South Korea) with Acridine Orange / Propidium Iodide stain (Nexcelom Bioscience, CAT# CS201065ML). Brightfield images of the samples were also taken using an EVOS-XL microscope (Advanced Microscopy Group; Bothell, Washington). A 500µL aliquot of washed saliva was removed and stored on ice as the “Whole” saliva fraction.

The remaining washed sample (~2.5 mL) was passed through a 30µm filter to capture large epithelial cells. Following filtration, the original collection tube was washed using 1mL BSA Rinsing Buffer, which was also passed through the filter. The filter was washed with an additional 1mL BSA Rinsing Buffer. The cells captured on the 30µm filter were rinsed off into a 50mL tube (Corning, CAT# 14-432-22), which we term the epithelial fraction, using 2mL BSA Rinsing Buffer. An additional 5mL BSA Rinsing Buffer was added to the epithelial fraction and the tube temporarily stored on ice.

The small cell filtrate was filtered through a second 30µm filter into a new tube to remove any remaining epithelial cells or cellular aggregates. The first filtrate tube and filter were washed again using 1mL BSA Rinsing Buffer each. The filtered small cells were prepared for magnetic bead antibody selection according to the Miltenyi instructions. We used magnetic separation with CD45 MicroBeads (Miltenyi, CAT# 130-045-801) to obtain one fraction of CD45+ immune cells and one of CD45-discard. The bead-sorted immune fractions were centrifuged at 350g for 5 minutes at 4°C. The whole and epithelial fractions were centrifuged at 500g for 5 minutes at 4°C. At the end of the sample processing, there were three fractions per sample: Whole, epithelial, and immune. In addition, there were six Oragene kit samples of whole saliva.

### Cell lysis, DNA extraction & quantitation, DNA methylation measurement

Large keratinocyte epithelial cells present in saliva were difficult to lyse and particular care was taken to prepare those cells. Whole, epithelial, immune, and Oragene fractions were resuspended in 500µL Buffer ATL (Qiagen, CAT# 69504). They were added to tissue disruptor bead tubes (MP Biomedicals, CAT# 116913100). The FastPrep-24 Tissue and Cell Homogenizer (MP Biomedicals; Irvine, CA) were used twice sequentially at a speed of 6.0 m/s for 35 seconds with the QuickPrep Adapter. The samples rested on ice for 5 minutes between shakes. To inactivate proteins, 20µL proteinase K (Qiagen, CAT# 69504) was added to all samples. Whole, epithelial, and immune fractions were incubated on a heat block for at least one hour at 56°C. The Oragene fractions were heated for two hours according to the DNA Genotek prepIT protocol.^22^

Genomic DNA was extracted from lysed cells using the DNeasy Blood & Tissue Kit (Qiagen, CAT# 69504) following the manufacturer’s protocol. DNA was eluted using two rounds of 50µL Buffer AE each. We processed 72 total fractions (22 whole, 22 epithelial, 22 immune, and 6 Oragene kits) for DNA extraction.

Nucleic acids were quantified using the NanoDrop 2000c (Thermo Scientific; Waltham, MA). Samples with a minimum of 250ng nucleic acid (22 whole, 22 epithelial, 22 immune, and 5 Oragene samples) were submitted to the Epigenomics Core at the University of Michigan for analysis using the Infinium MethylationEPIC BeadArray (Illumina, CAT# WG-317-1003). The Epigenomics Core used a Qubit 2.0 Fluorometer (Life Technologies; Carlsbad, CA) to measure the DNA concentration. Samples with a minimum 200ng DNA were used for DNA methylation measurements. To examine the reliability of the DNA methylation measurements of the epithelial cells, duplicates of six epithelial fractions were run as technical replicates. The 71 samples which passed the minimum 200ng DNA threshold were subjected to sodium bisulfite conversion and cleaning (EZ-96 DNA Methylation™ Kit, Zymo Research) according to the manufacturer’s instructions. Samples were then randomized and loaded on a single MethylationEPIC BeadArray plate. Fluorescence was measured using the iScan System (Illumina; San Diego, CA) at the Advanced Genomics Core at the University of Michigan.

### DNA methylation data preprocessing

EPIC BeadArray IDAT image files were processed and control metrics were assessed using the ewastools package.^23^ Background correction was performed using noob^24^ in the minfi package.^25^ Sex, predicted from the DNA methylation data, was compared to the survey demographic data. To ensure fractions derived from the same participant had the same genotype, SNPs were compared between fractions from each participant. Samples with >3% probes exceeding the detection p-value = 0.01 were dropped. For a preliminary estimate of the relative amounts of the epithelial and immune cell fractions in each sample, cell-type proportions were estimated for each sample using a reference panel generated from ENCODE and adult white blood cell data, implemented in ewastools. Based on this analysis, we excluded one epithelial cell fraction that was estimated to be >70% immune cells. In total, seven samples were excluded (control metrics n=1, sex comparison n=0, genotype comparison n=2, probe detection n=3, cell distribution n=1) (**Figure 1b**). To quantify the variation in measurements, the mean centered Pearson correlations of beta values (ratio between 0 and 1 of methylated and unmethylated alleles) were calculated for the five remaining epithelial technical replicate pairs (r=0.83, 0.88, 0.92, 0.94, 0.96). The paired sample from the technical replicates with the higher probe fail rate was excluded.

DNA methylation was measured at 866,091 sites. Following sample exclusion, probes with >5% of samples with detection p-values > 0.01 were dropped (n=10,298). Cross-reactive probes (n=43,254) and sex chromosome probes (n=19,681) were also dropped.^26,27^ The quality controlled DNA methylation data contained 795,694 probes from 18 epithelial, 20 immune, 18 whole saliva, and 4 Oragene samples (**Figure 1b**).

### Statistical and bioinformatic analysis

We calculated sample descriptive statistics on demographic and laboratory measures. For continuous variables (age, cell count, cell viability, sample volume), minimum, maximum median, and mean were calculated. For categorical variables (sex, race, illness status), count and frequency were provided.

To visualize DNA methylation distributions by cell type, density plots were constructed. To summarize variation in the DNA methylation data, principal component analysis was conducted. Principal components explaining cumulatively at least 90% of variance in the sample were examined. Principal components were tested for association with demographic and laboratory covariates using ANOVA tests for categorical variables and Pearson correlation tests for continuous variables.

To test for differences in DNA methylation between epithelial and immune cell samples, t-tests were used at each DNA methylation site. To adjust for multiple comparisons and identify probes that were significantly different by cell type, we used the Bonferroni significance level (p < 10^-8^). Among the significant probes, average methylation at each probe was calculated and plotted in a histogram. The 500 most statistically different probes by p-value were plotted in a heatmap with unbiased hierarchical clustering to group samples by similarity in probe profile. Global DNA methylation was calculated for each sample by averaging DNA methylation across all sites. Linear regression was used to test the association between the categorical variable, cell type (exposure), and global DNA methylation (outcome). We tested the 10,000 most statistically different probes for enrichment in gene ontology biological processes using the missMethyl package.^28^ Pathways with less than five annotated genes were excluded. Gene ontologies were constructed for both hypomethylated and hypermethylated probes.

To estimate saliva cell-type proportions in each sample using our saliva reference dataset, we integrated our new immune and epithelial sorted cell data into the ewastools package.^23^ Reference datasets are normalized and processed prior to package integration, which greatly reduces computation time for the end user, as well as provides more consistent cell proportion estimates across different datasets. Cell types were then estimated using the Houseman algorithm^9^ as applied by the *estimateLC* function.

To compare our new sorted saliva reference panel to datasets of similar cell types, we examined epithelial cell data from ENCODE and adult immune cell data.^29–31^ The ENCODE dataset contained DNA methylation data derived from eleven tissues around the body (**Supplemental Table 1**).^29,31^ The adult immune cell DNA methylation data was derived from seven magnetic bead sorted cell types (neutrophils, CD4+ T cells, CD8+ T cells, B cells, eosinophils, monocytes, and natural killer cells) donated by six men.^32^ We pre-processed the ENCODE and adult immune cell data using methods described above (**Supplemental Figure 1**). To understand the sample clustering, we conducted principal component analysis across all datasets.

To benchmark our new saliva DNA methylation cell-type reference dataset, we compared whole saliva DNA methylation-based cell-type proportions estimated using our new saliva reference dataset to those estimated using ENCODE epithelial data and adult immune cell data^29–31^ (**Supplemental Figure 2**). In whole saliva sample DNA methylation measures, we estimated cell-type proportions using both reference panels with the ewastools function *estimateLC*. Next, we calculated the variance explained by the estimated cell-type proportions using linear regression at each probe and calculated the R^2^ values. Across all probes, we calculated the R^2^ median as the average variance of DNA methylation values explained by estimated cell-type proportions.

As a sensitivity analysis, we compared the matched estimated cell proportions from Oragene and whole saliva samples from the same three people where both paired samples were available. To make a qualitative comparison of the reference panels, we estimated the cell proportions in the whole saliva samples using the ENCODE and adult white blood cell reference panel as well as our new saliva reference panel, both implemented in ewastools. To compare the epithelial and immune proportion estimates from each reference panel, we calculated a Pearson correlation for each cell type. We visualized the matched estimates with the sample brightfield image. As an exploratory analysis, we estimated specific immune cell types in the whole saliva samples, using the adult white blood cell reference panel,^32^ implemented in ewastools.^23^ The distribution of these immune cell types in whole saliva was then compared to the expected ranges in healthy pediatric peripheral blood.^33–38^

DNA methylation data are available through the Genome Expression Omnibus (GEO accession number GSE147318) and as a Bioconductor package (BeadSorted.Saliva.EPIC). Saliva cell-type estimation can be implemented through the ewastools package. All DNA methylation data preprocessing and analyses were conducted in R statistical software (version 3.6). Code to reproduce preprocessing and analyses is available (https://github.com/bakulskilab).

## Results

### Study sample description

Saliva samples were collected from 22 participants. One participant was excluded due to insufficient DNA for measurement in all fractions. Of the 21 participants with DNA methylation data, 15 were male and 10 were non-Hispanic white (**Table 1**). Three of the 21 participants were reported to be sick at the time of sample collection. The mean age was 11.8 years with a range of 7.9 – 16.9 years. From each participant, we collected a mean 3.1mL of unstimulated saliva. Collected saliva cell counts ranged from 720,000 to 34,000,000 cells per whole sample. Cell viability ranged from 5.1 – 81.5% and the median viability was 69.2%. Following microscopic evaluation, large, flat, and geometric epithelial cells were easily differentiated from the small, round immune cells (**Figure 2**). These microscopy images highlight the interindividual heterogeneity in cell size, shape, and proportions in saliva samples.

**Table 1.** Descriptive statistics for participants (n=21). Saliva was collected and 18 whole saliva samples, 18 epithelial fractions, 20 immune fractions, and 4 Oragene kit whole samples passed DNA methylation quality control measures.

|  | Mean (sd) | Count (%) | Range |
| --- | --- | --- | --- |
| Sex (Male) |  | 15 (71.4) |  |
| Race |  |  |  |
| White |  | 10 (47.6) |  |
| Black |  | 1 (4.8) |  |
| Biracial* |  | 10 (47.6) |  |
| Sick |  | 3 (14.3) |  |
| Age (years) | 11.8 (2.7) |  | (7.9, 16.9) |
| Cell Count<sup>+</sup> | $6.3 \times 10^6$ ($9.8 \times 10^6$) | | ($7.2 \times 10^5$, $3.4 \times 10^7$) |
| Viability (%)<sup>+</sup> | 69.2 (16.7) |  | (5.1, 81.5) |
| Volume (mL)<sup>+</sup> | 3.5 (1.4) |  | (1.9, 6.5) |

\* Asian/White or Other/White
<sup>+</sup>Median used due to non-normal distribution

**Figure 2.**
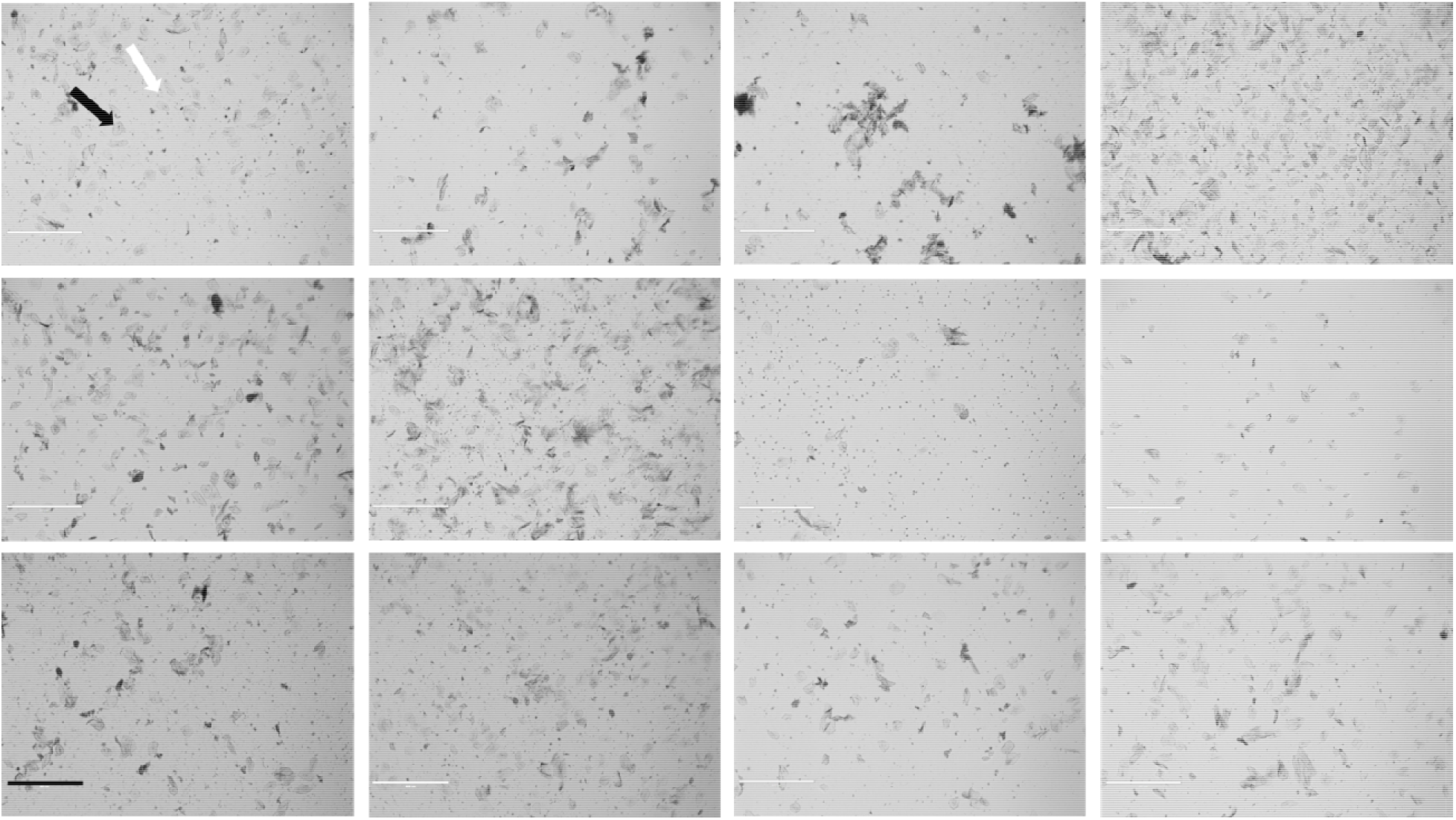
Representative sample of saliva diversity from 12 participants. Images were taken using a brightfield microscope at 4x (EVOS-xl) following the resuspension in BSA Rinsing Buffer. The black arrow points to an epithelial cell and the white arrow points to an immune cell. Scale bar = 400μm.

### Assessment of differences in DNA methylation between saliva cell types

Following quality control, DNA methylation from 18 epithelial cell fractions, 20 immune cell fractions, 18 whole samples, and four Oragene kits were included in this analysis. A total of 795,694 probes were analyzed (**Figure 1b**). The mean global DNA methylation of the immune cells was 57.5% (standard error: 0.2%). The mean global DNA methylation of epithelial cells was 53.2% (standard error: 0.2%) (**Supplemental Figure 3**). Immune cells had 4.3% (standard error: 0.3%) higher mean global methylation compared to the epithelial cells (p = 2 × 10^-16^). The density plot reflected the expected bimodal DNA methylation distribution as measured by probes (**Figure 3a**). A principal component analysis of the immune and epithelial cell DNA methylation data showed that the first principal component of the DNA methylation data explained 80.8% of the variance and was associated with cell type (p = 1.7 × 10^-15^) (**Figure 3b**). The second principle component explained 4.6% of the variance in the DNA methylation data and was associated with participant age (p = 0.004), saliva sample cell viability (p = 0.004), and participant sex (p = 0.03).

**Figure 3.**
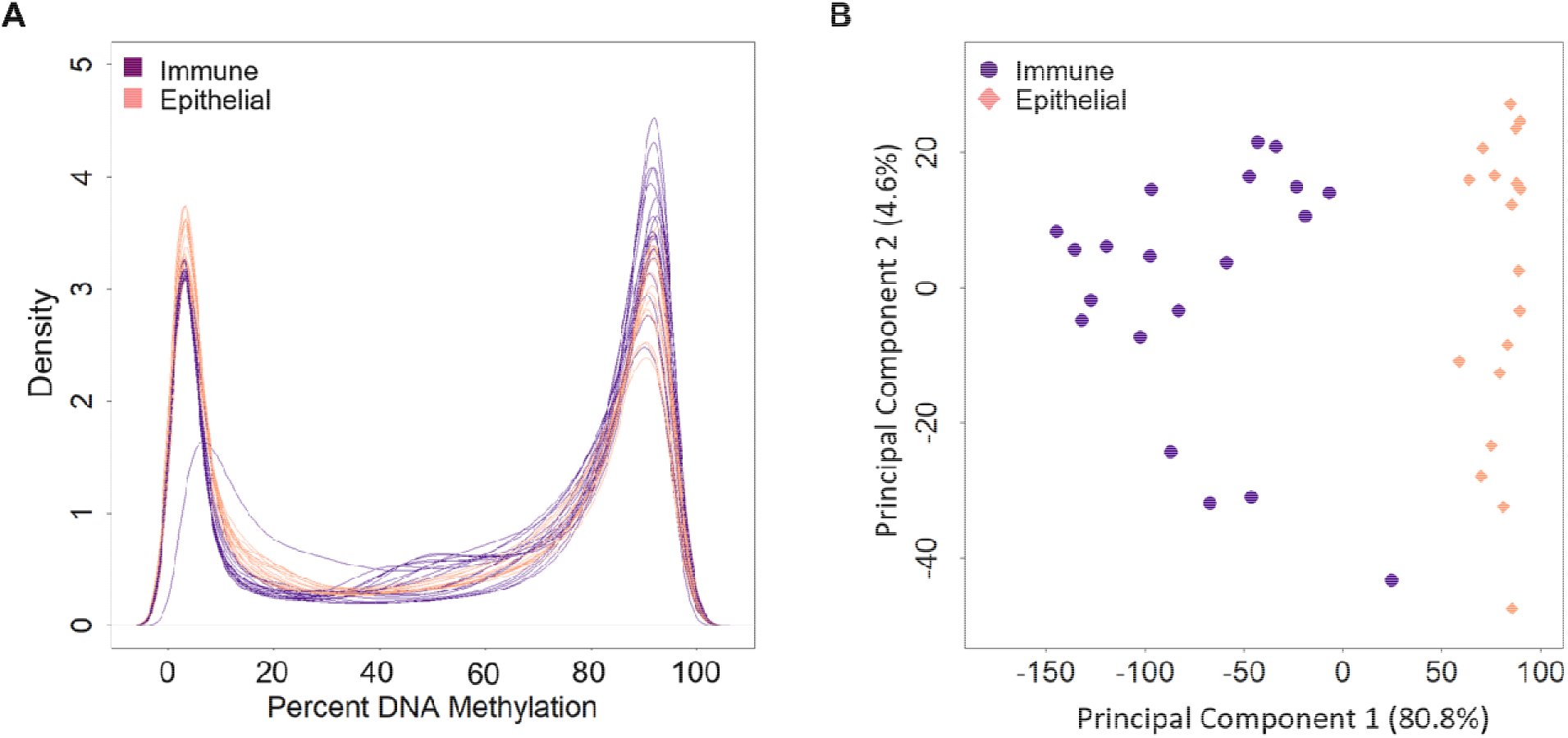
Density plot and principal component analysis of DNA methylation measures painted cell types. **A)** Density plots of DNA methylation of all CpG sites, displaying DNA methylation distributions by cell type (immune cells in purple and epithelial cells in pink). Each line represents one cell fraction. Beta values were converted to percentages. **B)** DNA methylation principal components one and two, colored by immune cells in purple and epithelial cells in pink. Principal component 1 explained 80.8% of the variance in the DNA methylation data. Principal component 2 explained 4.6% of the variance in the DNA methylation data.

DNA methylation levels at each site were compared between epithelial and immune cells using t-tests. We identified 164,793 (20.7% of all probes) differentially methylated sites between epithelial and immune cells (p < 10^-8^). Among the genome-wide significantly differentially methylated sites, 72.2% had higher DNA methylation in immune cells relative to epithelial cells (**Figure 4a**). Among the differentially methylated sites, the average magnitude of DNA methylation difference was 32.4%. 27.8% of sites were hypomethylated in immune cells relative to epithelial cells. The highest magnitude differences were observed at cg07110356 in the MPO gene (Myeloperoxidase) with 70.2% higher methylation in epithelial cells compared to immune cells (p = 8.2 × 10^-18^) (**Supplemental Table 2**) and at cg17804342 in RGS10 gene (Regulator of G Protein Signaling 10) with 68.6% higher DNA methylation in immune cells compared to epithelial cells (p = 7.6 × 10^-20^). The 500 most statistically differentially methylated sites between immune and epithelial cells were analyzed by unbiased hierarchical clustering and visualized by heatmap (**Figure 4b**). Saliva sample fractions clustered by cell type. Among these 500 sites, 93.6% had higher DNA methylation in immune cells relative to the epithelial cells (**Supplemental Table 2**).

**Figure 4.**
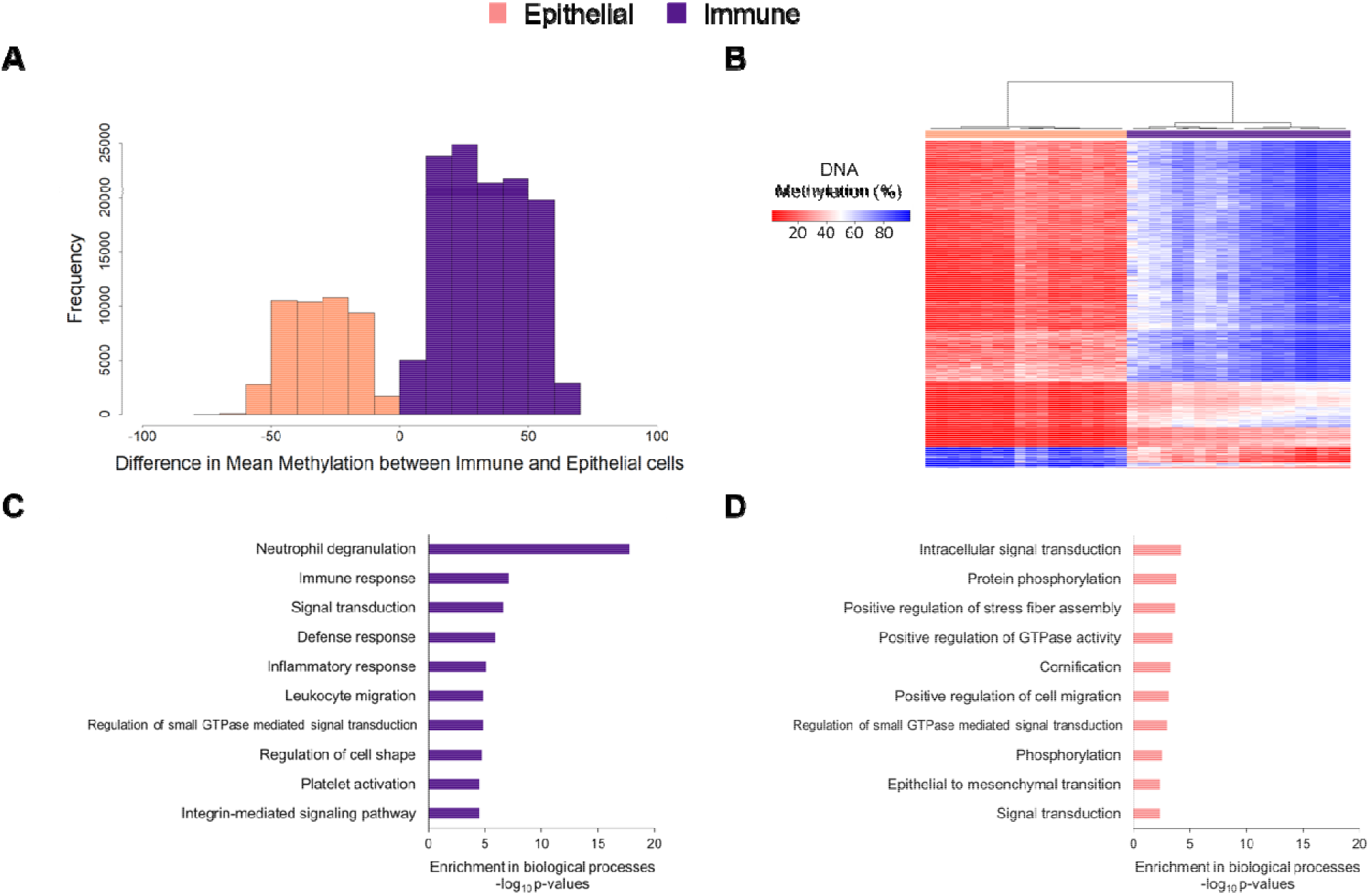
DNA methylation differences between immune and epithelial cell types. **A)** Among DNA methylation sites differentially methylated between immune and epithelial cells (p < 10^-8^, 164,793 sites), histogram of the magnitude of DNA methylation difference. The x-axis is percent nt methylation, and the reference group was epithelial cells. The values were calculated as percent methylation of immune minus that of epithelial. **B)** The top 500 most differentially methylated sites between immune and epithelial cells by p-value (t-test) are plotted in heatmap rows (red indicates lower DNA methylation, blue indicates higher DNA methylation). In heatmap columns,, unbiased hierarchical clustering of samples was performed (immune fractions in purple and epithelial fractions in pink). **C)** Bar chart of the gene ontology biological processes enriched (minimum p < 3.2 × 10^-5^) among genes hypomethylated in immune cells, relative to epithelial cells. **D)** Barchart of the gene ontology biological processes enriched (minimum p < 4.7 × 10^-3^) among genes hypomethylated in epithelial cells, relative to immune cells.

We tested for enriched gene ontology biological pathways in the differentially methylated sites between immune and epithelial cells. Sites with lower DNA methylation in immune cells mapped to genes enriched for immune pathways such as neutrophil degranulation (p = 1.6×10^-18^), immune response (p = 7.6×10^-8^), and leukocyte migration (p = 1.5×10^-5^) (**Figure 4c**). Sites with lower DNA methylation in epithelial cells mapped to general cell activity pathways such as intracellular signal transduction (p = 6.3×10^-5^), protein phosphorylation (p = 1.6×10^-4^), and positive regulation of stress fiber assembly (p = 2.2×10^-4^) (**Figure 4d**). Cornification, a key process for hard palate formation, was also enriched for differentially methylated genes in epithelial cells (p = 5.2×10^-4^).

### Cell proportion estimation

DNA methylation data from epithelial cell types in ENCODE and primary adult immune samples were used to estimate saliva cell proportions. DNA methylation data from these samples together with our saliva samples analyzed by principal component analysis revealed samples primarily clustered by cell type and study (**Supplemental Figure 4**). Principal component 1 explained 47.9% of the variation in the data and was associated with cell type (p = 1.8 × 10^-50^) and study (p = 4.3 × 10^-27^) variable. Principal component 2 explained 16.1% of the variation in the data and was also associated with cell type (p = 2.3 × 10^-51^) and study (p = 6.1 × 10^-48^). We first estimated cell proportions in our saliva DNA methylation data using a reference panel constructed from ENCODE epithelial cells and adult white blood cells, implemented in ewastools.^29–31^ Our saliva epithelial cell fractions were estimated to be 81.8 – 97.4% epithelial cells (median: 93.4%, IQR: 4.7%) and the immune cell fractions were 13.0 – 88.9% immune cells (median: 58.9%, IQR: 26.4%) (**Figure 5a**). Our whole saliva samples were estimated to be 8.4 - 88.3% epithelial cells (median: 71.8%, IQR: 25.6%) and 10.8 – 88.1% immune cells (median: 29.9%, IQR: 24.6%) (**Supplemental Figure 5a)**. Our Oragene saliva samples were estimated to be 19.8 – 59.8% epithelial cells (median: 28.5%) and 30.6 – 69.6% immune cells (median: 55.4%). Estimated cell percentages derived from the ENCODE and adult white blood cell reference panel explained a median of 33.6% of the variation in the whole saliva DNA methylation data (**Supplemental Figure 6**).

**Figure 5.**
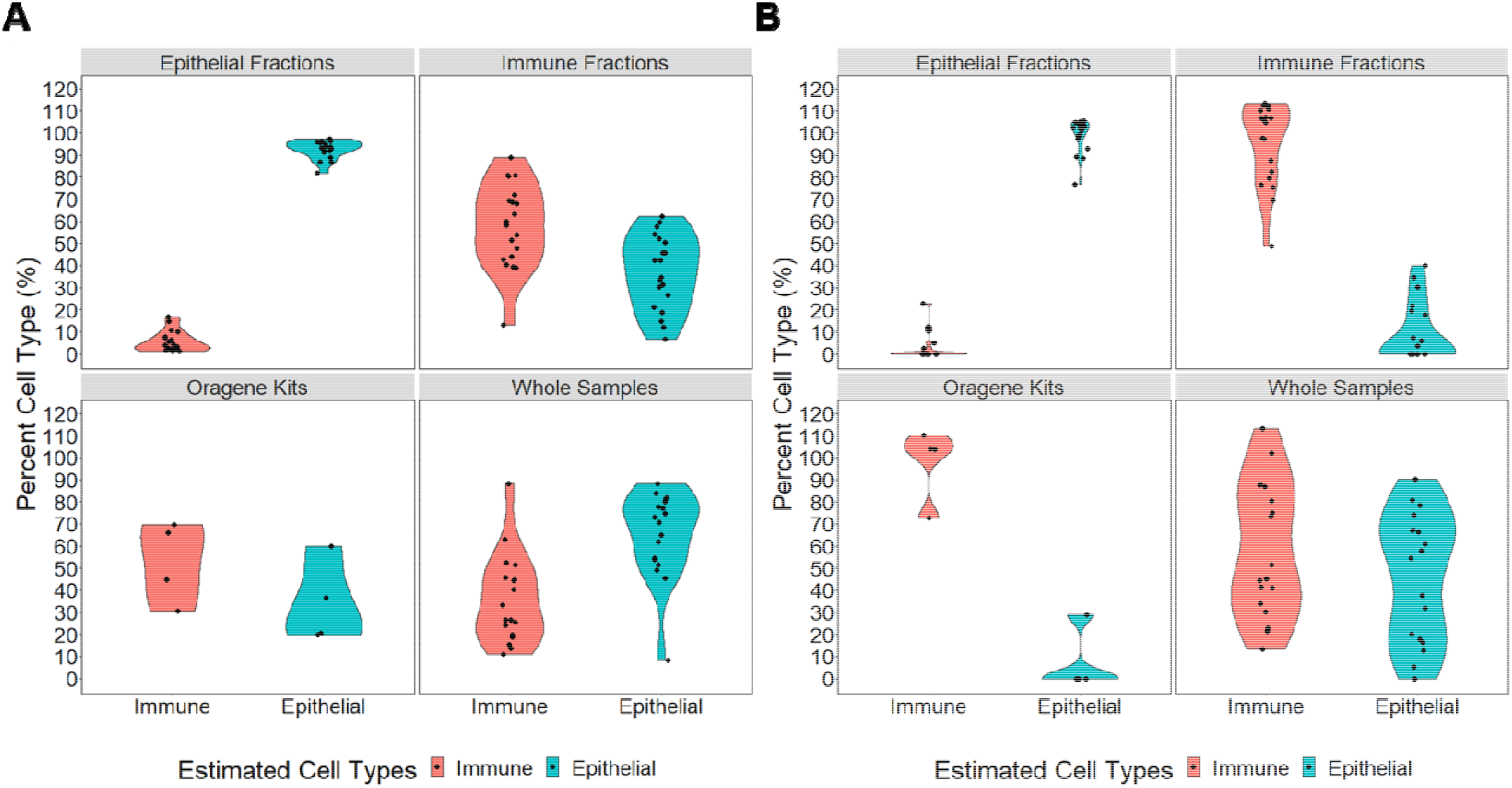
Cell-type percentages estimated in saliva samples from DNA methylation data, using two estimation methods. Violin plots are used to visualize the percent immune cells estimated in red and the percent epithelial cells estimated in blue. In both panels A and B, the upper left quadrant shows the percent cell types estimated in sorted epithelial samples (n=18); the upper right quadrant shows the percent cell types estimated in sorted immune cell samples (n=20), the lower left quadrant shows the percent cell types estimated in whole saliva collected in Oragene kits (n=4); and the lower right quadrant shows the percent cell types estimated in whole saliva samples collected directly (n=18). **A)** Percent cell types were estimated using a reference panel generated from ENCODE epithelial cell DNA methylation data^29,31^ and an adult white blood cell data,^30^ implemented through ewastools. **B)** Percent cell types estimated using our new primary saliva reference panel implemented through ewastools.^23^

We next estimated cell proportions in our saliva DNA methylation data using our new saliva reference data integrated into the ewastools package. The new primary saliva reference panel-derived cell proportions explained a median of 31.5% of the variation in the whole saliva sample DNA methylation data. Our saliva epithelial cell fractions were estimated to be 76.6% – 105.3% epithelial cells (median: 102.2%, IQR: 6.1%) (**Figure 5b**). Saliva immune cell fractions were 48.9% – 113.5% immune cells (median: 105.4%, IQR: 28.6%). Our whole saliva samples were estimated to be 0 – 90.3% epithelial cells (median: 56.1%, IQR: 48.8%) and 13.2 – 113.2% immune cells (median: 48.2%, IQR: 49.5%) (**Supplemental Figure 5b**). Our Oragene saliva samples were estimated to be 0.0 – 28.9% epithelial cells (median: 0.0%) and 72.8 – 110.0% immune cells (median: 103.8%). Whole saliva samples that had a high predicted proportion of immune cells correspondingly had a low predicted proportion of epithelial cells. For example, whole saliva sample 19 had the highest predicted immune proportion and the lowest epithelial predicted proportion.

In an exploratory analysis, using our new saliva reference panel, we compared the estimated immune cell proportions in three Oragene saliva samples to three matched whole saliva samples from the same participants. The Oragene saliva samples had an average of 27.7% higher estimated immune cell proportions compared to the whole saliva samples (**Supplemental Figure 5c**).

We compared whole saliva DNA methylation cell proportions estimated using the ENCODE epithelial and adult immune reference panel to cell proportions estimated using our new primary saliva reference panel. Immune cell proportions estimated by the two methods were highly correlated (r = 0.96) (**Supplemental Figure 7a**). The new saliva reference panel estimated higher mean immune proportion (58.4%) compared to the ENCODE epithelial and adult immune cell reference panel (34.2%). Epithelial cell proportions estimated by the two methods were also highly correlated (r = 0.91) (**Supplemental Figure 7b**). The new saliva reference panel estimated lower mean epithelial proportion (46.6%) compared to the ENCODE epithelial and adult immune cell reference panel (65.6%). At the extremes, both models were less linear. We visually compared the whole saliva cell proportions estimated from both reference panels to the brightfield images of the whole saliva samples (**Supplemental Figure 8**).

We estimated immune cell subtypes in saliva using the adult white blood cell reference panel^32^ and compared the proportions to literature ranges of immune cells in pediatric peripheral blood (no standard saliva cell proportions were available). In the saliva immune cell fractions, Oragene samples, and whole samples, the range of estimated granulocytes was wider than the normal range in blood (**Supplemental Figure 9**). Saliva immune fractions were estimated to be 30 – 100% granulocytes, while in pediatric blood, the normal range was 38 – 72%. The estimated range of granulocytes in the Oragene samples was 41 – 86%. The estimated range of granulocytes in the whole samples was 20 – 100%. Saliva estimated CD4+ T-cell proportions had a similar range to pediatric blood, though saliva estimates were lower. For example, saliva immune fractions were estimated to be 0 – 28% CD4+ T-cells, while in pediatric blood the normal range was 31 – 52%. In all saliva samples, no CD8+ T-cells were estimated, though the normal range of CD8+ T-cells in pediatric blood was 18 – 35%. In general, the estimated ranges of immune cell proportions in saliva was more variable than in blood.

## Discussion

Saliva is a commonly used biosample for epigenetic epidemiology studies, and especially in pediatric studies. A saliva-specific cell-type reference panel was critically needed to estimate cell-type proportions from bulk saliva DNA methylation data in children. This gap was particularly salient in light of the substantial interindividual heterogeneity in salivary cell-type composition (highlighted in the microscopy images in **Figure 2**). We collected whole saliva samples and Oragene kits from children and sorted the whole samples into immune and epithelial fractions based on a combination of size and antibody-based sorting, and the DNA methylation profiles of each were measured using the Illumina MethylationEPIC BeadChip. We identified substantial differences in DNA methylation patterns between the sorted cell fractions with sites enriched for logical biological pathways (e.g. immune pathways in the immune fraction, cornification in the epithelial fraction). Our datasets were integrated into the publicly available ewastools^23^ and produced a saliva reference panel R data package (BeadSorted.Saliva.EPIC)^39^ to facilitate cell-type proportion estimation. Future saliva DNA methylation studies will be able to easily integrate this reference panel for cell-proportion estimation into their current analytic workflow.

We compared the performance of our saliva reference panel to a reference panel we generated using ENCODE epithelial cells and adult immune cell data. There was a strong, positive correlation (immune cells: r = 0.96, epithelial cells: r = 0.91) between the whole saliva cell proportion estimates using our reference panel and estimates in the same samples using the ENCODE epithelial cells and adult white blood cells reference panel. The ENCODE and adult white blood cell reference panel, on average, explained slightly more of the variance in saliva DNA methylation data compared to our saliva reference panel, implemented in ewastools. Because the surface-level oral mucosa only contains a few types of epithelial cells,^17^ the higher variance explained by the ENCODE and adult white blood cell reference panel could be a result of the larger number and range of epithelial cell types included from ENCODE. Unfortunately, there is no existing external validation dataset of cell counts in saliva to compare our new saliva reference panel to the ENCODE epithelial and adult immune cell reference panel data. However, using the new saliva reference panel, we observed a more dynamic range in the cell-type proportions estimated in whole saliva samples (**Supplemental Figure 5a-b**), which is consistent with the highly variable biosamples (**Supplemental Figure 8**). We further recommend the use of the new saliva reference panel for pediatric saliva epigenetic studies, as reference panels based on primary site-specific tissues may be more biologically relevant.

There was substantial interindividual variability in the proportions of cell types in saliva. We estimated our whole samples to have an average of 46.6% epithelial cells (**Figure 5b**), with an interquartile range of 49.5%. Similarly, another study found an interquartile range of epithelial cells saliva in children to be 46.3%, and the cell-type variability in saliva was higher in children compared to adults.^40^ Large interindividual differences in saliva sample cell proportions can drive the DNA methylation profile and therefore influence results.^20^ In this study, we observed cumulative cell percentages exceeding 100% in whole saliva, which is consistent with the recommended ewastools cell-type implementation using unconstrained estimates to explain greater variance in DNA methylation.^23^ Together, these findings emphasize the importance of understanding the proportions of cell types in DNA methylation analyses, especially when using saliva.

We observed that saliva from children had wider ranges of granulocytes, CD4+ T cells, and monocytes compared to the normal blood ranges for children. Granulocytes had the highest estimated proportion of immune cells in our saliva samples, similar to a study that manually counted segmented immune cells (granulocytes) in saliva from children.^40^ No CD8+ T-cells were predicted in any saliva sample from the present study (**Supplemental Figure 9**). Using flow cytometry to sort saliva immune cells from three participants, T-cells ranged from 0.8% to 1.2%, but they did not separate out CD8+ cells.^16^ Both CD4+ and CD8+ T-cells have been identified in salivary glands^41^ which suggests that CD8+ T-cells are present in oral cavity tissue, though they may not migrate into saliva. Variability observed in saliva cell types from relatively healthy children could influence observed differences in DNA methylation between groups.

Many large saliva epigenetic studies use Oragene kits to collect biosamples. Oragene kits provide long storage time and high DNA quality and yield.^42^ In our small comparison (n=3) of matched whole samples collected in an empty tube and samples collected in Oragene kits, we estimated higher immune cell proportions in the Oragene kit samples. These preliminary findings suggest that the Oragene kits may enrich for immune cell DNA. In our experience, keratinized saliva epithelial cells are considerably more resistant to lysis than immune cells. It is possible that epithelial cells are inadequately lysed, enriching for immune cell DNA, and potentially biasing DNA methylation measures. A larger and specifically designed study is needed to explore this trend.

Our study had a number of limitations. We isolated two main saliva cell types: epithelial and immune cells. In each of these types, there were likely several subtypes of cells that we grouped into one category. For example, there are several subtypes of epithelial cells that cover the oral surfaces.^17^ Papanicolaou staining of buccal samples from children have identified three main epithelial cell types: intermediate squamous, non-keratinous, and keratinous superficial squamous.^40^ Surface markers for flow cytometry sorting are not well characterized for normal oral epithelial cells. We initially attempted to isolate epithelial cells using flow cytometry with an antibody for epithelial cell adhesion molecule (EpCAM), a typical surface marker on epithelial cells,^43–45^ but oral epithelial cells do not appear to express EpCAM (data not shown). In addition, we found that the large size of saliva epithelial cells, which can be up to 100µm in diameter, blocked the microfluidic tubes of the flow cytometry and droplet-based single cell instruments. We also attempted nuclear isolation for single nuclei sequencing, but the recommended detergent was insufficient to lyse the epithelial cells, possibly due to their highly keratinized nature. Future studies may use other methods to isolate and profile different epithelial populations. Although we combined saliva immune cells into one category as well, there are existing reference panels from blood that can be used to predict leukocyte proportions.^32^

Our study also has a number of strengths. While our participants were not a random sample, we included saliva samples from 21 children with 18 epithelial cell samples and 20 immune cell samples. We identified 164,793 significantly different (p < 10^-8^) DNA methylation sites between epithelial and immune cells, highlighting that the separation procedure isolated biologically distinct cellular populations. For comparison, the commonly used adult blood reference panel based on six participants, observed 37,837 sites that differentiated at least one leukocyte cell type.^10,30^ We also included five technical replicates of epithelial cell fractions. The average mean centered correlation between the replicates was 0.91. The epithelial fractions, immune fractions, whole samples, and Oragene kit samples were randomized on the slides and all run on one plate to reduce batch effects. Using our new saliva reference panel, saliva cell proportion estimation improved on currently available methods while maintaining a high degree of association (**Supplemental Figure 7**). This is the first primary saliva reference panel for cell-type proportion deconvolution in saliva. This saliva reference panel will improve epigenetic studies by providing a tool that is both appropriate and easy to use for estimating cell-type proportions for use in regression modeling approaches.

### Conclusions

DNA methylation differences measured in saliva are likely influenced by the relative proportions of cells present in the sample.^46^ Epithelial and immune cells from saliva have distinct DNA methylation profiles, and there is substantial interindividual heterogeneity in the cell proportions in saliva. Changes in the proportion of epithelial to immune cells will influence whole saliva DNA methylation measurements. Our reference panel and accompanying R package (BeadSorted.Saliva.EPIC)^39^ provide a new, more biologically relevant, method to better account for cell-type proportions in pediatric saliva DNA methylation research, which is an important step to account for cell-type effects in epigenetic studies.

## Data Availability

DNA methylation data are available through the Genome Expression Omnibus (GEO accession number GSE147318) and as a Bioconductor package (BeadSorted.Saliva.EPIC). Code to reproduce preprocessing and analyses is available (https://github.com/bakulskilab).

https://www.ncbi.nlm.nih.gov/geo/query/acc.cgi?acc=GSE147318

## Funding and Acknowledgements

This study was approved under the University of Michigan Institutional Review Board (HUM00154853). We thank the University of Michigan Epigenomics Core and Advanced Genomics Core for DNA methylation measures. Support for this research was provided by the National Institute of Environmental Health Sciences (grants P30 ES017885, R01 ES028802, U01 ES026697, R35 ES031686, R01 ES025531, R01 ES025574), the National Institute on Aging (grants R01 AG067592, R01 AG060110-01), the National Institute on Minority Health and Health Disparities (grants R01 MD011716, R01 MD013299), and the National Institutes of Health Office of the Director (grants UG3 OD023285, UH3 OD023285).

